# High Altitude Protects Against Early Childhood Obesity: Nationwide Evidence from Colombia

**DOI:** 10.1101/2025.07.14.25329127

**Authors:** Fernando Lizcano, Eliana Avilés, Álvaro Javier Burgos-Cárdenas, Daniela Sanabria, Lizeth Montañez, Fredy Luna, Miguel O’Mehara, Alex Valenzuela

## Abstract

Childhood obesity is an emerging public health challenge globally, with early-life exposures and environmental factors critically shaping long-term metabolic risk. Among these, high-altitude hypoxia has been proposed to influence energy metabolism and adiposity regulation; however, its relationship with early childhood obesity remains insufficiently explored at a population level. This ecological study examined the association between residential altitude and obesity prevalence in children under five years of age across Colombia.

Data were obtained from 1,123 municipalities using official national databases: obesity prevalence from the Colombian Institute of Family Welfare (ICBF), population estimates from the National Department of Statistics (DANE), and altitude data from official geographic registries. Municipalities were stratified into four altitude ranges and categorized by population size. We calculated obesity prevalence and odds ratios (OR) to evaluate the risk associated with altitude, adjusting for urbanicity and demographic scale.

Among approximately 4.16 million children aged 0–5 years, those residing between 2,000 and 3,000 meters above sea level had significantly lower odds of obesity compared to those living below 1,000 meters (OR: 0.47). This inverse association remained consistent in departmental capital cities and in municipalities stratified by similar population sizes. Notably, higher obesity prevalence in small, low-altitude municipalities suggested potential deficiencies in healthcare access and early prevention.

These findings support the hypothesis that altitude may serve as an environmental modulator of early metabolic risk, aligning with the developmental origins of health and disease (DOHaD) framework. Our results call for geographically tailored public health strategies, integrating biological and structural determinants to address early-onset obesity. Further longitudinal and mechanistic studies are warranted to better understand the pathways linking hypoxia, metabolic regulation, and childhood obesity.

## Introduction

Childhood obesity has become one of the most serious public health challenges of the 21st century. According to the World Health Organization, in 2022, more than 39 million children under the age of 5 were overweight or obese worldwide, with a high economic impact ^1–3^. In Latin America, childhood overweight rates have risen sharply in the past two decades, with early childhood (0-5 years) being a particularly vulnerable window for the establishment of long-term metabolic programming ^4^. In Colombia, national nutrition surveys have shown increasing trends in early childhood overweight and obesity, disproportionately affecting urban and low-income populations^5^. Obesity in early life results from a complex interplay of genetic, epigenetic, environmental, and behavioral factors ^6^. At the physiological level, key contributors include altered appetite regulation, impaired energy expenditure, and insulin resistance^7^ . Mitochondrial dysfunction has also been implicated, leading to inefficient oxidative phosphorylation and increased lipid storage ^8^ . Dysregulation of adipose tissue signaling, including leptin and adiponectin pathways, may further disrupt energy homeostasis^9^ . In infants and toddlers, these disturbances can be exacerbated by early-life exposures, such as maternal overnutrition, gestational diabetes, and suboptimal breastfeeding practices^10–12^.

Exposure to high-altitude environments (typically above 2000 meters above sea level) introduces a hypobaric hypoxic condition that affects oxygen delivery, metabolic rate, and systemic physiology^13^ . Several studies have hypothesized that altitude may exert protective metabolic effects, including increased basal energy expenditure, altered appetite-regulating hormones (e.g., ghrelin, leptin), and upregulation of genes associated with oxidative metabolism^14^ . However, an increase in insulin resistance may reduce the favorable effect of high altitude ^15^ . Research in adults from Peru, Bolivia, and Tibet has shown a lower prevalence of obesity and metabolic syndrome at higher altitudes. However, similar evidence in children, particularly those under 2 years old ^6^ ^16^ ^17^, is scarce and contradictory. A constellation of maternal, environmental, and developmental factors influences the risk of obesity in early childhood. Studies in Latin American countries such as Ecuador, Peru, and Argentina have reported conflicting findings regarding the association between altitude, obesity, and growth in children aged 0–5 years^18^. In Ecuador, research comparing children residing in urban highland areas (2,772 meters) with those in coastal regions (605 meters) found that highland children exhibited lower height-for-age and weight-for-age Z-scores, with stunting and underweight rates increasing by factors of four and three, respectively ^19^ .

In Peru, studies have utilized an altitude threshold of 2,500 meters to differentiate settings. One study found that living at altitudes below 2,500 meters was associated with a 2.67-fold higher likelihood of obesity. Additional reports highlighted significant regional disparities: obesity prevalence reached 10.1% in more urbanized areas like Lima, while it dropped to 2.6% in rural jungle regions^20^.

In Argentina, children residing at altitudes above 2,500 meters were found to be shorter and lighter than their counterparts in low-lying areas. Still, they had higher body mass index (BMI), ponderal index, and surface area-to-body mass ratios^21^. Age-specific analyses revealed that stunting was most pronounced before 24 months of age, and obesity rates were elevated during the first year of life. The socioeconomic context further influenced these patterns: urban environments at higher altitudes with lower poverty rates were associated with higher obesity prevalence, whereas poverty in highland areas was linked to chronic malnutrition^22^.

Multiple risk factors, including maternal obesity and metabolic status during pregnancy, quality of prenatal nutrition, maternal education, socioeconomic level, breastfeeding duration, and exposure to obesogenic environments, may influence obesity in early childhood. The “developmental origins of health and disease” (DOHaD) hypothesis suggests that intrauterine and early postnatal exposures program long-term metabolic outcomes^23^ ^24^. Thus, investigating how these risk factors interact with altitude may help to explain observed geographical disparities in obesity prevalence.

In this ecological study^25^, we analyzed data from 1,123 municipalities across Colombia to explore whether altitude is associated with the prevalence of obesity in children under five years of age. Using national health surveillance data and population estimates, municipalities were stratified into four altitude ranges (<1,000 m; 1,000–1,999 m; 2,000–2,499 m; and ≥2,500 m). We found that the prevalence of childhood obesity was highest in low-altitude municipalities (≥6%) and decreased consistently with elevation, reaching a minimum in regions above 2,500 meters. This trend remained significant after stratifying by population size and examining department capitals. Our findings suggest that living at high altitude may confer a protective effect against early childhood obesity, likely mediated by both physiological adaptations and socioeconomic gradients.

## Materials and Methods

Study design and data sources. This study was designed as a cross-sectional ecological analysis based on secondary data from official public health and demographic sources. We included all 1,123 municipalities in Colombia with available data on childhood obesity, estimated population, and altitude.

### Data sources

We obtained childhood obesity data from the Colombian Institute for Family Welfare (Instituto Colombiano de Bienestar Familiar, ICBF), based on the Sistema de Información *Cuéntame* - *Tomás Nutricionales*-^26^. This system compiles data from ICBF’s regional offices throughout the country and includes nutritional assessments of children under the agency’s care. These assessments are conducted by health professionals trained in anthropometric measurement, following the official ICBF manual. The data are considered highly reliable, as they are used for clinical purposes to detect children at risk of malnutrition. The reporting protocol is standardized across all regions of Colombia, and the ICBF currently provides services to approximately 1.4 million children.

For this study, the ICBF provided aggregated data at the municipal level on the number of children aged 0–5 years diagnosed with obesity, based on standardized weight-for-height tables for early childhood. The data cut-off was October 31, 2024. To estimate population-level obesity rates, we utilized municipal-level population projections from the National Administrative Department of Statistics (Departamento Administrativo Nacional de Estadística, DANE in Spanish) (https://www.dane.gov.co/.), specifically those for children aged 0-5 years in 2024. These denominators enabled standardized comparisons of obesity prevalence across municipalities, considering differences in population size.

To further account for socioeconomic disparities between municipalities, we incorporated the Unmet Basic Needs Index (NBI, Necesidades Básicas Insatisfechas in Spanish), a composite poverty indicator developed by DANE using data from Colombia’s most recent National Population and Housing Census, conducted in 2018. The NBI captures the proportion of individuals experiencing at least one of the following deprivations: extreme poverty, inadequate housing, lack of access to basic public services, household overcrowding, school absenteeism, and high economic dependency. It is recognized as a reliable and valid measure and is widely used by the Colombian government to guide and implement public policy. For each municipality, the NBI was included as a continuous covariate in a multivariable logistic regression model to evaluate its association with obesity prevalence and to control for potential confounding due to structural socioeconomic inequalities. For altitude data, we searched the Official altitudes of municipal seats provided by the Instituto Geográfico Agustín Codazzi (IGAC). https://www.igac.gov.co/.

### Statistical analysis

All statistical analyses were conducted using a combination of open-source and licensed platforms to ensure reproducibility and analytical robustness. Data processing, cleaning, and integration were performed using Python 3.10 with the Pandas, NumPy, and Matplotlib libraries. Stratification by altitude and population size, as well as data visualization, were developed using the Seaborn and Matplotlib packages.

For univariate analyses, we calculated proportions, prevalence, odds, and 95% confidence intervals using binomial approximation (Wilson method). Chi-square tests for independence were applied to compare proportions of obesity across altitude strata. These tests were executed using the SciPy stats and stats models libraries in Python.

Multivariate stratification (e.g., by capital city status and population group) was conducted through cross-tabulation and stratified subgroup analyses. Although no formal regression model was applied, odds ratios (OR) were estimated using classical formulas: OR = (a/c) / (b/d), with 95% confidence intervals derived from the log-normal approximation for relative risk estimations.

Geographic integration was based on altitude ranges obtained from municipal seat data (IGAC), and all municipal identifiers were normalized for consistency across datasets. Data validation steps included cross-matching administrative codes from DANE and ICBF, Cuéntame System, with altitude records from the National Geographic Institute. No imputation of missing data was performed. All analyses were conducted in a local environment using macOS and Ubuntu Linux, with Python, and figures were rendered in high-resolution vector formats suitable for publication.

The prevalence of obesity was calculated for each municipality by dividing the number of reported cases by the estimated population of children aged 0-5 years. Municipalities were grouped into four altitude categories: 0-1000, 1001-2000, 2001-3000, and >3000 meters above sea level (m.a.s.l.). Additionally, municipalities were stratified into five categories based on their child population size: less than 1,000, 1,000-5,000, 5,001-10,000, 10,001-20,000, and more than 20,000. The capital cities of Colombia’s 32 departments were flagged as a separate analytic subgroup.

Descriptive statistics were used to summarize child population size, number of obesity cases, and prevalence across altitude and municipality size categories. To evaluate the association between altitude and obesity, we calculated odds ratios (ORs), using the 0-1000 m.a.s.l. The odds of obesity were calculated as p / (1 − p), where p is the prevalence in each stratum of altitude. Chi-square tests were used to assess differences in the odds of obesity across altitude groups. The category serves as the reference group to estimate the relative likelihood of obesity in higher altitude strata.

Stratified analyses were conducted in capital cities and towns with fewer than 5,000 inhabitants to control for potential differences in healthcare access and reporting practices. All data were anonymized and aggregated, and the analysis was conducted in accordance with ethical standards for the secondary use of public health surveillance data.

A multivariable logistic regression analysis was also performed to estimate the association between municipal obesity prevalence and altitude, adjusting for three indicators from the Unmet Basic Needs Index (NBI): overall NBI score, the proportion of the population in extreme poverty, and the housing quality component. The outcome variable was dichotomized, with municipalities classified as having a high prevalence of obesity if the rate exceeded 1%. We proceed with a quadratic transformation for altitude in a model to detect U shape effect.

## Results

The sample consisted of data from 1,123 municipalities across Colombia. Stratified by altitude, the first stratum (0-1000 meters above sea level) included 471 municipalities, with a total population of 2,314,464 children aged 0-5 years, and showed an obesity prevalence of 0.84%. The second stratum (1001-2000 m.a.s.l.) encompassed 341 municipalities and 897,058 children, with a prevalence of 0.80%. In the third stratum (2001-3000 m.a.s.l.), 181 municipalities and 1,005,106 children were recorded, with a notably lower prevalence of 0.40%. The fourth stratum (>3000 m.a.s.l.), with only 7 municipalities and 11,498 children, presented a prevalence of 0.86%.

Compared with the reference group (0-1000 m), the odds ratio (OR) of childhood obesity was 0.95 in the second stratum, 0.47 in the third, and 1.03 in the fourth. These differences suggest a protective effect associated with living at elevations of 2001-3000 m above sea level. (Table 1). The altitude-related variation in obesity prevalence is visualized in Figure 1.

**Figure 1.**
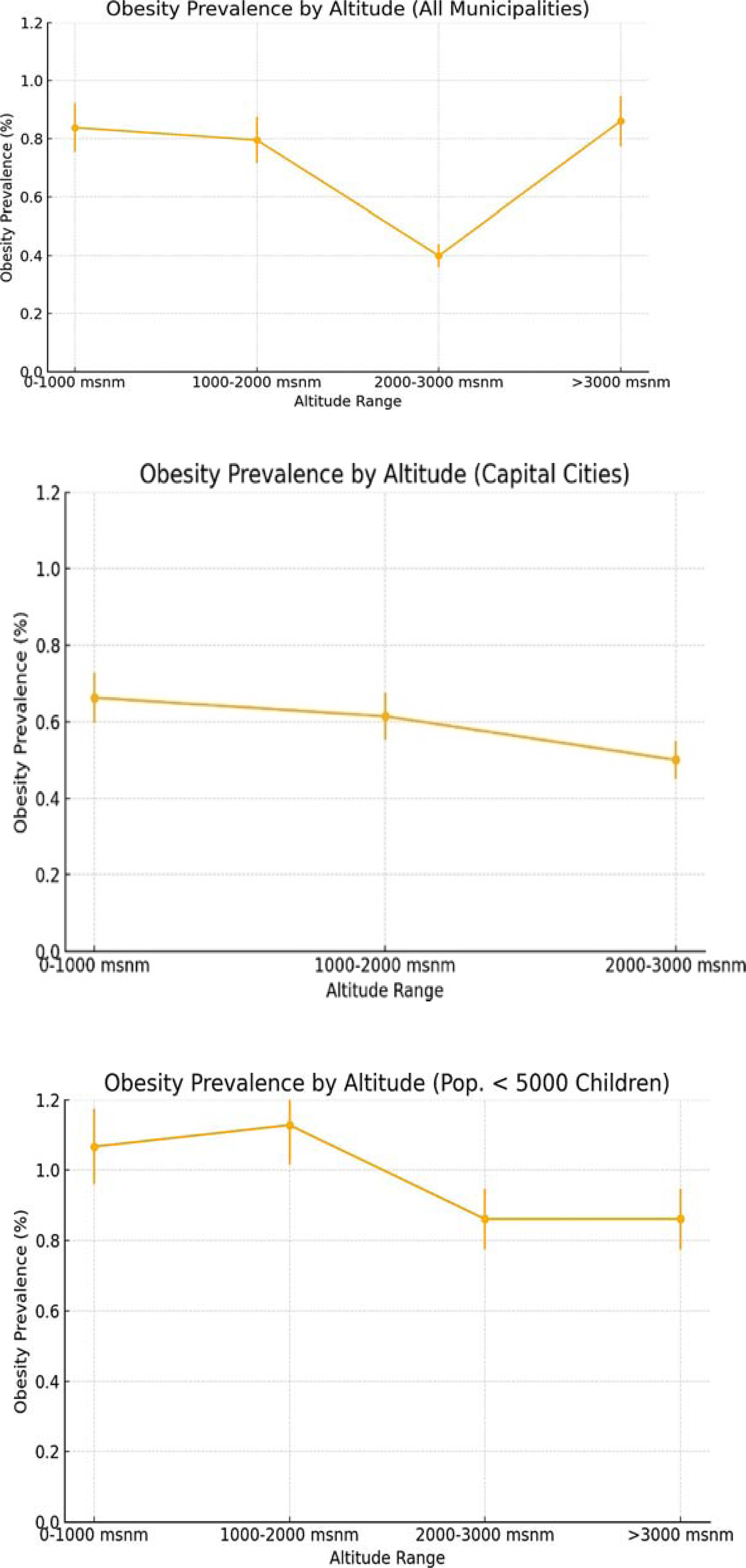
Obesity prevalence by altitude in children aged 0-5 years. The superior plot shows the prevalence according to altitude in all municipalities. The middle plot shows the prevalence according to altitude in capital cities, and the lower plot in municipalities with less than 5000 children. The data in the middle plot were taken under the assumption that childcare was of higher quality in the departmental capitals compared to other smaller municipalities, where primary care may be less effective. The chart in the lower evaluates municipalities with inferior populations to assess the circumstances in more marginalized regions of the country with less support in health services or basic needs. These factors can be confounding when evaluating the risk of altitude and obesity in early childhood.

**Table 1.**
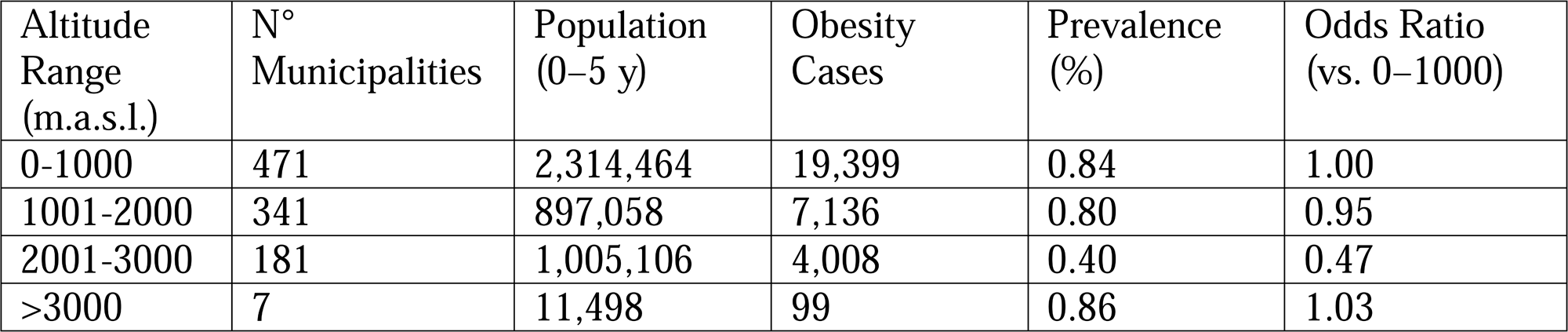
Childhood Obesity Prevalence by Altitude Strata in Colombian Municipalities.

### Stratified analysis

The prevalence of obesity calculated in capital departmental cities and small municipalities with less than 5,000 children between 0-5 years had a similar tendency that the total sample fig 1. the prevalence of obesity was highest in areas below 1000 meters and progressively lower at higher altitudes, with the lowest observed in municipalities between 2000-3000 meters (0.40%). The univariate OR shows a similar tendency in capitals and municipalities with fewer than 5,000 children (see Table 1).

### Multivariate and Stratified Analyses

The altitude was associated with a higher probability of having 1 percent prevalence of obesity after adjusting for NBI index, extreme poverty, and housing composition. Regarding socioeconomics variables, the NBI index and extreme poverty were associated with the probability of having a prevalence greater than 1%. The estimator, p-value and calibration measured are showed in table 2.

**Table 2.**
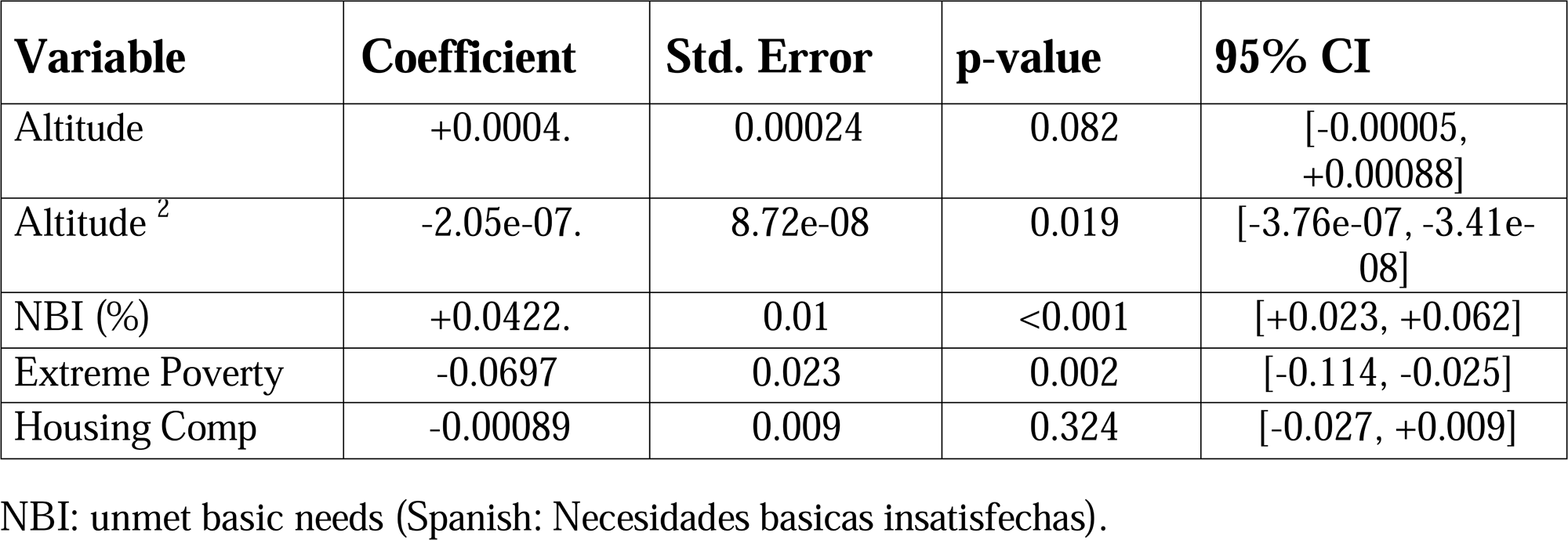
Coefficients from the multivariate logistic regression model predicting elevated obesity risk.

The model was statistically significant overall (p < 0.001; pseudo-R² = 0.018). Altitude showed a non-linear relationship with obesity. The linear term was positive (p = 0.082), while the quadratic term was negative and significant (p = 0.019), indicating an inverted U-shaped relationship, where the risk of obesity decreases at higher altitudes is visualized in Figure 2. NBI was positively associated with obesity (p < 0.001), while extreme poverty was negatively associated (p = 0.002). Housing quality was not significantly associated.

**Figure 2.**
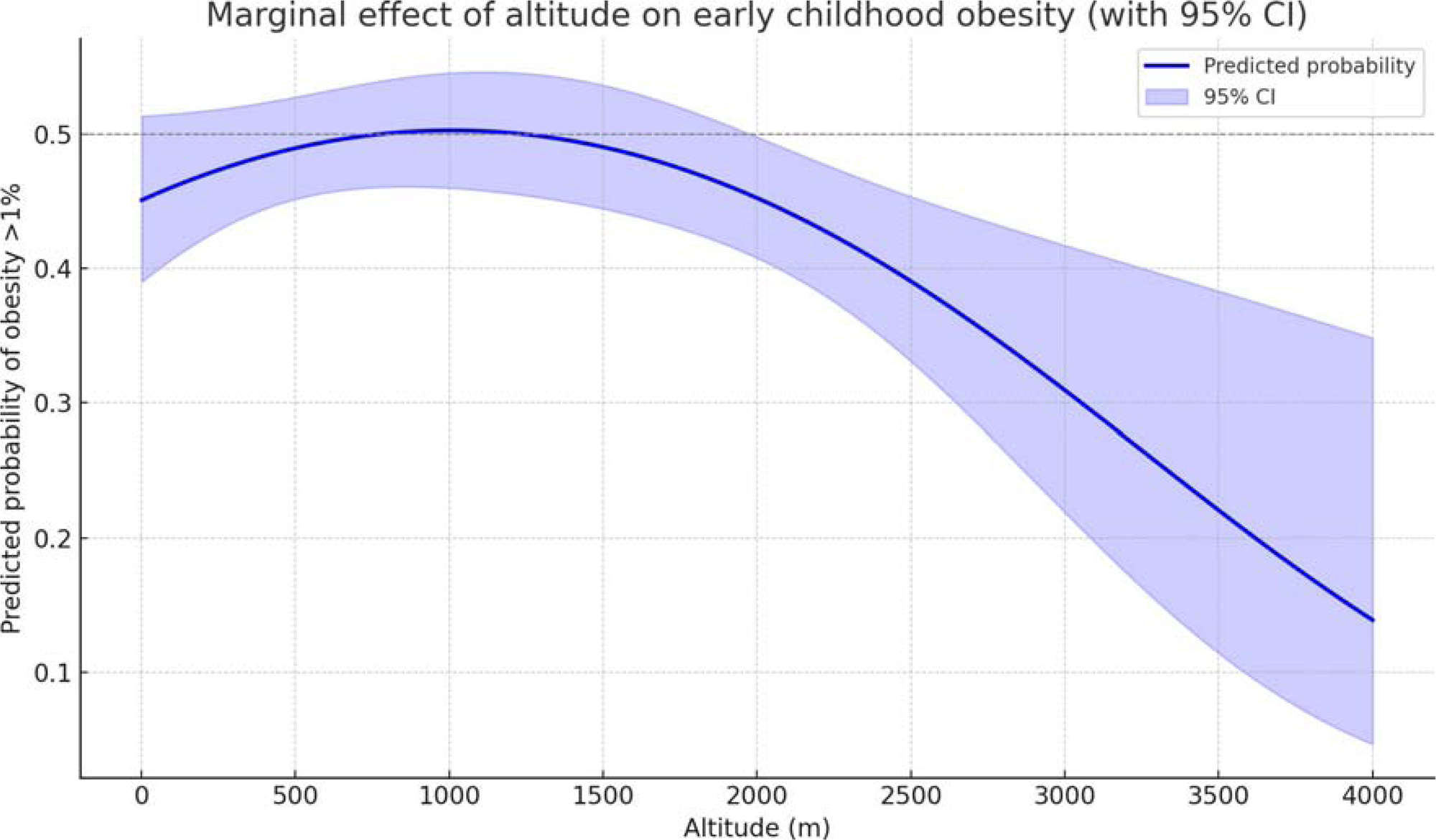
Predicted probability of early childhood obesity (>1%) with altitude, based on a multivariate logistic regression model. Altitude was included as both a linear and quadratic term, adjusted for the average values of three socioeconomic indicators: proportion of people with unmet basic needs (NBI), proportion of the population in extreme poverty, and a housing quality index. The figure illustrates a non-linear inverse relationship between altitude and obesity, with a pronounced decline in predicted probability above ∼1,500 meters above sea level. The shaded region represents the 95% confidence interval. The slight increase above ∼3,500 meters should be interpreted cautiously, as it may reflect sparse data, high environmental stress, or urban transitions in a few extreme-altitude communities.

A stratified model was also run in municipalities with more than 5,000 children. In this subgroup (n = 140), the protective effect of altitude was stronger and statistically significant (p = 0.011), and the quadratic term was not significant. The pseudo R ² improved substantially (from 0.179), suggesting a better model fit in larger, possibly better-equipped municipalities, visualized in Figure 3.

**Figure 3.**
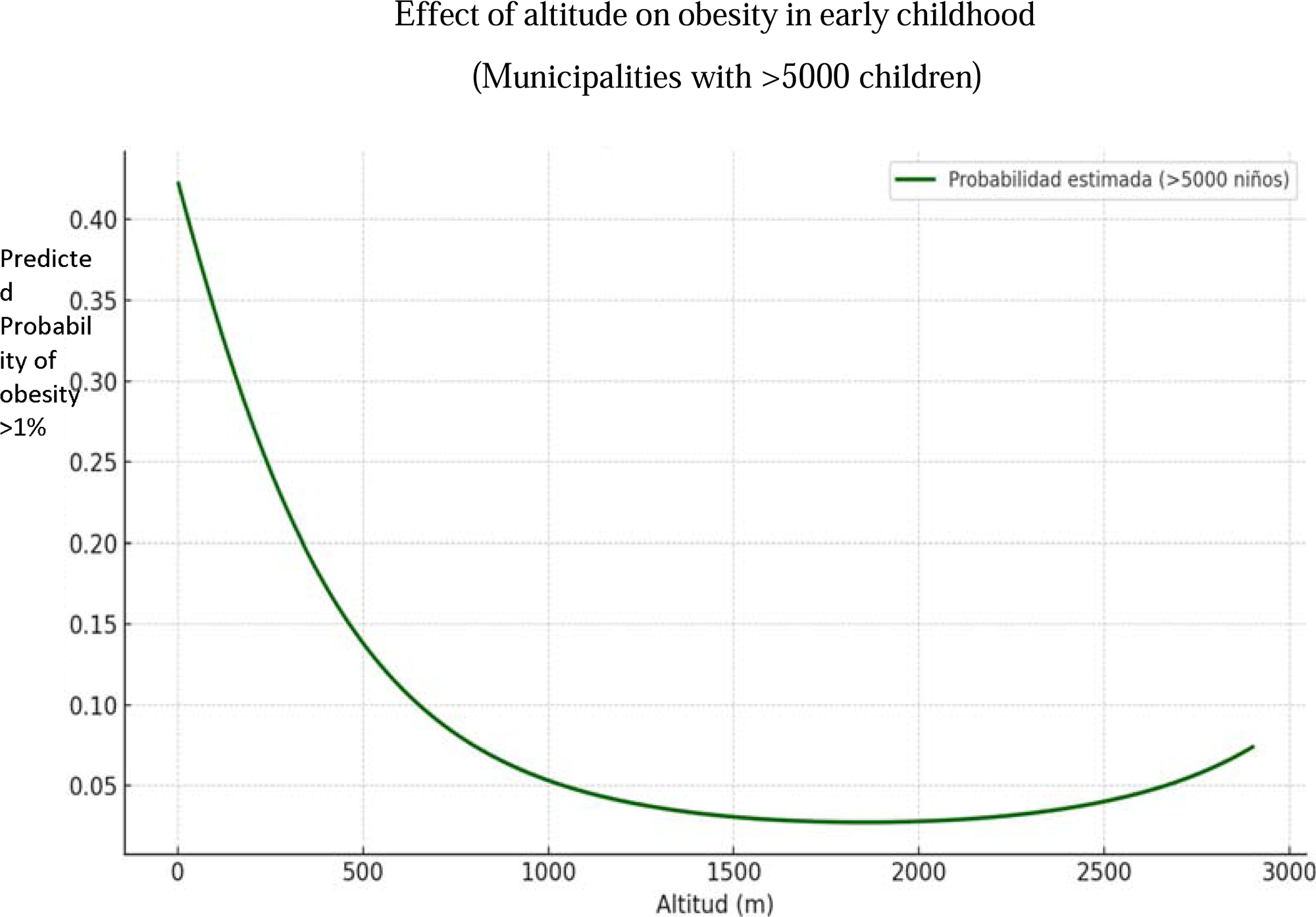
Estimated probability of elevated obesity in early childhood by altitude, in municipalities with a child population lower than 5,000. Curve generated from a multivariate logistic model adjusted for the proportion of people with unmet basic needs, poverty, and housing conditions. A clear linear downward relationship is observed, with altitude acting as a significant protective factor against childhood obesity. In this subgroup, which primarily includes capital cities or large urban centers, the model explained a greater proportion of the variance (pseudo-R ² = 0.179).

These findings support the hypothesis that altitude acts as a protective factor against obesity, particularly in more affluent areas, and that underlying socioeconomic conditions may influence this effect.

## Discussion and Conclusions

This study provides evidence that residing at higher altitudes (>2,000 meters above sea level) is associated with a reduced prevalence of early childhood obesity in Colombia, even after adjusting for socioeconomic indicators, including unmet basic needs and extreme poverty. The multivariate analysis revealed a significant inverse relationship between altitude and obesity rates among children aged 0-5 years, suggesting that environmental factors linked to altitude may confer protective effects against excessive weight gain in early life.

Multiple epidemiologic studies in high-altitude regions of Latin America consistently show an inverse association between altitude and childhood obesity. In the Peruvian Andes, for example, Santos et al.^27^ reported that the prevalence of overweight among youth (6-16 years) was significantly lower in high-altitude areas (∼6% in the high Andes) compared to low-altitude coastal regions (∼41% at sea level).

Similarly, a Bolivian study comparing schoolchildren from a high-altitude rural district (∼4000 m) with those from a low-altitude subtropical village (∼650 m) revealed that only 8% of the high-altitude children were overweight or obese, compared to 41% in the low-altitude setting^28^ . These patterns strongly suggest that, as altitude increases, the prevalence of childhood obesity tends to decrease. Evidence from the Himalayas supports this trend: in the Tibetan Highlands, childhood overweight and obesity rates are significantly lower than national averages. A survey of 7- to 12-year-old children in the Tibetan highlands (with an average altitude of approximately 4,500 m) revealed an overweight prevalence of only 8.5% (with obesity at 6.3%), which is significantly lower than that among children in lowland China^29^.

Overall, studies from the Andes (Peru, Bolivia, Ecuador, and Argentina) and the highlands of Asia indicate a strong negative correlation between altitude and the risk of childhood obesity, with populations at higher altitudes generally being thinner than those at lower altitudes. Notably, multilevel analyses in Peru confirm that, even after controlling for individual and school-related factors, living at high altitude independently predicts a lower likelihood of childhood overweight^30^.

Several physiological mechanisms may induce this association. In adolescents and adults, high-altitude environments are characterized by hypobaric hypoxia, which can lead to increased basal metabolic rates and altered hormonal regulation of appetite^31^. Studies have shown that hypoxia-inducible factors (HIFs) activated at high altitudes can upregulate leptin expression, thereby improve leptin sensitivity and contributing to appetite suppression and energy expenditure^8^ ^32^. Additionally, exposure to cold temperatures prevalent at higher elevations may further elevate energy expenditure through thermogenesis. These adaptations may collectively contribute to lower body mass indices observed in high-altitude populations^33^. However, the relationship between altitude and weight is complex and influenced by numerous factors during early infancy. While higher altitude is linked to lower birth weight and slower growth in infants, there is no substantial evidence that altitude acts as a protective factor against early infantile obesity. The primary effect of altitude appears to be growth restriction, rather than the prevention of excess weight or obesity in infancy^34^.

Our findings highlight a divergent influence of socioeconomic indicators on early childhood obesity. While the proportion of people living with NBI was positively associated with obesity prevalence, likely due to poor dietary quality and increased consumption of low-cost, ultra-processed foods, extreme poverty showed an inverse relationship. This contrast supports the ’double burden of malnutrition’ hypothesis: children in moderate poverty may be at risk for obesity due to poor diet, while those in extreme deprivation may suffer from caloric and protein undernutrition, which lowers the likelihood of excess weight^35^. These findings underscore the need to distinguish between different forms of socioeconomic vulnerability when designing interventions.

The findings highlight the intricate relationship between environmental and socioeconomic factors in the development of early childhood obesity. While altitude-related hypoxia appears to offer a protective effect, socioeconomic challenges can negate these benefits, highlighting the need for comprehensive public health strategies that address both environmental and social determinants of health

## Limitations

First, the ecological design does not allow inference at the individual level. Second, the dataset may be affected by underreporting in rural areas or places with limited healthcare access. Third, although socioeconomic indicators were included, unmeasured confounders such as maternal obesity, diet quality, or local health policies may influence the outcomes. Finally, altitude was measured based on the main municipal seat, which may not fully reflect conditions in geographically extensive municipalities.

In Colombia, national nutrition surveys have indicated a rising trend in early childhood overweight and obesity, disproportionately impacting urban and low-income populations. The etiology of early-life obesity is multifactorial, involving genetic, epigenetic, environmental, and behavioral components. Physiologically, key contributors include dysregulated appetite control, impaired energy expenditure, insulin resistance, and mitochondrial dysfunction, which can lead to inefficient oxidative phosphorylation and increased lipid accumulation. Additionally, disruptions in adipose tissue signaling pathways, such as those involving leptin and adiponectin, may further compromise energy homeostasis. Early-life exposures, including maternal overnutrition, gestational diabetes, and inadequate breastfeeding practices often exacerbate these disturbances

## Data Availability

The data from this study are Ecological data. no are data form individual personas.

https://www.dane.gov.co/

https://www.igac.gov.co/

